# Differences between meeting 24-hour movement guidelines and physical fitness levels among Japanese elementary school children

**DOI:** 10.1101/2024.12.14.24319036

**Authors:** Takashi Naito, Kenryu Aoyagi, Koichiro Oka, Kaori Ishii

## Abstract

Current physical fitness levels in Japanese children are lower than that in the 1980s. Investigating the specific relationship between adherence to 24-hour movement guidelines (24-h MG, MVPA: moderate-to-vigorous physical activity; ScT: screen time; Sleep: sleep duration) and physical fitness is crucial to improve children’s fitness levels. Overall physical fitness and eight physical fitness component scores (handgrip strength, sit-up, sit-and-reach, repeated side jump, 20-meter shuttle run, 50-meter sprint, standing long jump, and softball throw) were measured using standardized tests in Japanese elementary schools. Associations between compliance with 24-h MG, overall physical fitness, and each physical fitness component score were analyzed by analysis of covariance. A total of 307 participants (41.4% male) were included in the analysis. Overall fitness scores were significantly higher in children who met only MVPA (P < 0.01), MVPA and ScT (P < 0.01), MVPA and Sleep (P < 0.001), and all three criteria (P < 0.05), than in those who did not. Children meeting only MVPA, MVPA and ScT, and MVPA and Sleep criteria, had higher scores for physical fitness components than those who did not. Children who met one or both of ScT and Sleep criteria, did not differ significantly for overall fitness or its components scores compared to those who did not meet. Handgrip strength and flexibility were not associated with any 24-h MG pattern. Furthermore, compliance with MVPA combined with other behaviors (ScT and Sleep) demonstrated a trend towards greater overall fitness, muscle strength in lower extremities, cardiorespiratory fitness, agility, and dexterity.

## Introduction

Previous research has revealed that 81% of children and adolescents worldwide are physically inactive[1] and spend an excessive portion of their time in a sedentary manner [2,3]. These lifestyles may have partially contributed to the observed decline in physical fitness of children over the past few decades [4–8]. Similarly, the physical fitness of students in Japanese elementary schools has been declining since 1985 [9,10]. The factors underlying this trend include a reduction in the time spent performing physical activity and outdoor play in daily life[11] and a revision of the Guidelines for the Course of Study, which reduced physical education classes in elementary schools and shifted the focus of classes from physical fitness to emphasize enjoyment [12]. The development of physical fitness in childhood is critical because the level of physical fitness is associated with physical and mental health in childhood [13,14], along with adolescent mental health and cognitive performance [15].

Previous studies revealed an association between the levels of physical fitness in children and a range of lifestyle behaviors, including physical activity, sedentary behavior, and sleep [13,16–19]. The concept of 24-hour movement guidelines (24-h MG), which aim to achieve these three movement behavior recommendations in a comprehensive manner rather than individually, has become increasingly widespread [20–24]. 24-h MG encompasses the movement behaviors of physical activity, sedentary behavior, and sleep, over a 24-h period during which these behaviors complement each other. In Canada and Australia, 24-h MG is recommended for children aged 5–17 years, as follows: (1) 60 minutes of daily moderate-to-vigorous physical activity (MVPA), (2) no more than 2 hours of screen time (ScT) per day, and (3) 9–11 hours (ages 5–13 years) and 8–10 hours (ages 14–17) years of sleep duration (Sleep) [20,25].

Several previous studies have revealed associations between compliance with 24-h MG and physical fitness, with more compliance items of the three 24-h MG behaviors associated with greater aerobic capacity, muscular strength, and overall fitness [26–28]. Behavioral adherence to all possible combinations of MVPA, ScT and Sleep have also been investigated [26–29]. A systematic review revealed that adherence to MVPA in 24-h MG was especially associated with higher levels of physical fitness [30]. However, the number of studies involving children remains limited, and only few studies have investigated the relationship between each physical fitness component, such as muscle strength, cardiorespiratory fitness, agility, and dexterity.

In Japan, the physical fitness of children has declined since 1985 [10], although this change is not consistent across all measurement categories of physical fitness. Thus, investigating the association between 24-h MG and individual physical fitness factors, as well as overall physical fitness, is critical when considering measures to improve the physical fitness of children. Previous studies have also shown that adherence to 24-h MG is associated with obesity in children [31,32], mental health [33,34], self-reported health status [35], brain development [36], and academic achievement [37–39]. Therefore, application of 24-h MG has the potential to improve physical, mental, and cognitive health, in addition to physical fitness, thereby improving the overall well-being of children.

The purpose of this study was to comprehensively investigate the relationship between 24-h MG and overall physical fitness scores, and each physical fitness component score using data collected from an eight-event physical fitness test[40] that is standardized and implemented in elementary schools in Japan.

## Materials and Methods

### Participants

This study was conducted with a cohort of children attending six elementary school grades (1-6) in Japan. A total of 477 children (222 boys and 255 girls) aged 6-11 years in 2021 were included. Written informed consent to participate in the study was obtained from both the parents and children. This study was approved by Waseda University Ethics Review Procedures Concerning Research with Human Subjects (approval number: 2024-HN032). All studies were performed in accordance with relevant guidelines and regulations. For children in the first to third grades of elementary school, parents completed the survey form on their behalf; for children in the fourth to sixth grades, the children themselves responded to the survey form.

### MVPA survey

A questionnaire was used to assess the number of days in a typical week on which participants engaged in MVPA for 60 min or more per day. Using a validated questionnaire [41,42], MVPA was defined to include all such increases in breathing activities, such as walking, running, brisk walking, bicycle riding, dancing, swimming, soccer, and basketball, as well as physical education classes and recess activities. Children who performed physical activity for at least 60 min per day on 7 days per week were defined as meeting 24-h MG criteria, and those who performed physical activity for 6 days or less per week were defined as not meeting the criteria.

### Survey of screen time

A questionnaire was used to survey screen time in three situations: (1) watching television or videos, (2) playing video games, and (3) using the internet or e-mail outside of class time [43]. Participants were asked to indicate the number of days, and the average time duration per day they viewed or used each of these situations on weekdays and weekends. The weekday and weekend screen times were summed and divided by seven to calculate the mean screen time per day. Children with a screen time of ≤ 2 h per day were classified as meeting 24-h MG criteria, while those with > 2 h were classified as not meeting the criteria.

### Survey of sleep duration

A questionnaire was administered to determine the number of hours slept per day. The respondents were asked to rate their sleep duration using the following three options: (1) < 9 h, (2) 9–11 h, and (3) ≧ 11 h. Participants who responded 9–11 h were classified as meeting 24-h MG criteria, while those who responded with < 9 h and ≧ 11 h were classified as not meeting the criteria.

### Measurement of physical fitness

Physical fitness test scores were collated from students in elementary school. The physical fitness assessment comprised eight components: handgrip strength as a measure of muscle strength, sit-up as an indicator of trunk muscle strength and endurance, sit-and-reach as a measure of flexibility, repetitive side jump as an assessment of agility, a 20-m shuttle run as an evaluation of cardiorespiratory fitness, a 50-m sprint as a test of speed, a standing long jump as a measure of lower-limb muscle strength, and a softball throw as an assessment of explosive power and dexterity. Each component was evaluated on a 10-point scale (1–10) utilizing standardized criteria based on performance. Then, we calculated the score for each component and summated them to get the total score (8–80).

### Statistical analysis

Analysis of covariance (ANCOVA) was used to investigate the association between compliance with 24-h MG and physical fitness scores. The covariates were gender, grade, BMI, calculated height, and weight (in the year 2021) and compliance with MVPA, ScT, and Sleep criteria. The significance level was set at P < 0.05. All statistical analyses were performed using IBM SPSS (version 29.0; IBM Corp., Armonk, NY, USA).

## Results

Table 1 summarizes the characteristics of the participants. A total of 307 participants (41.4% male) were included in our final analysis after excluding those with missing data.

**Table 1.**
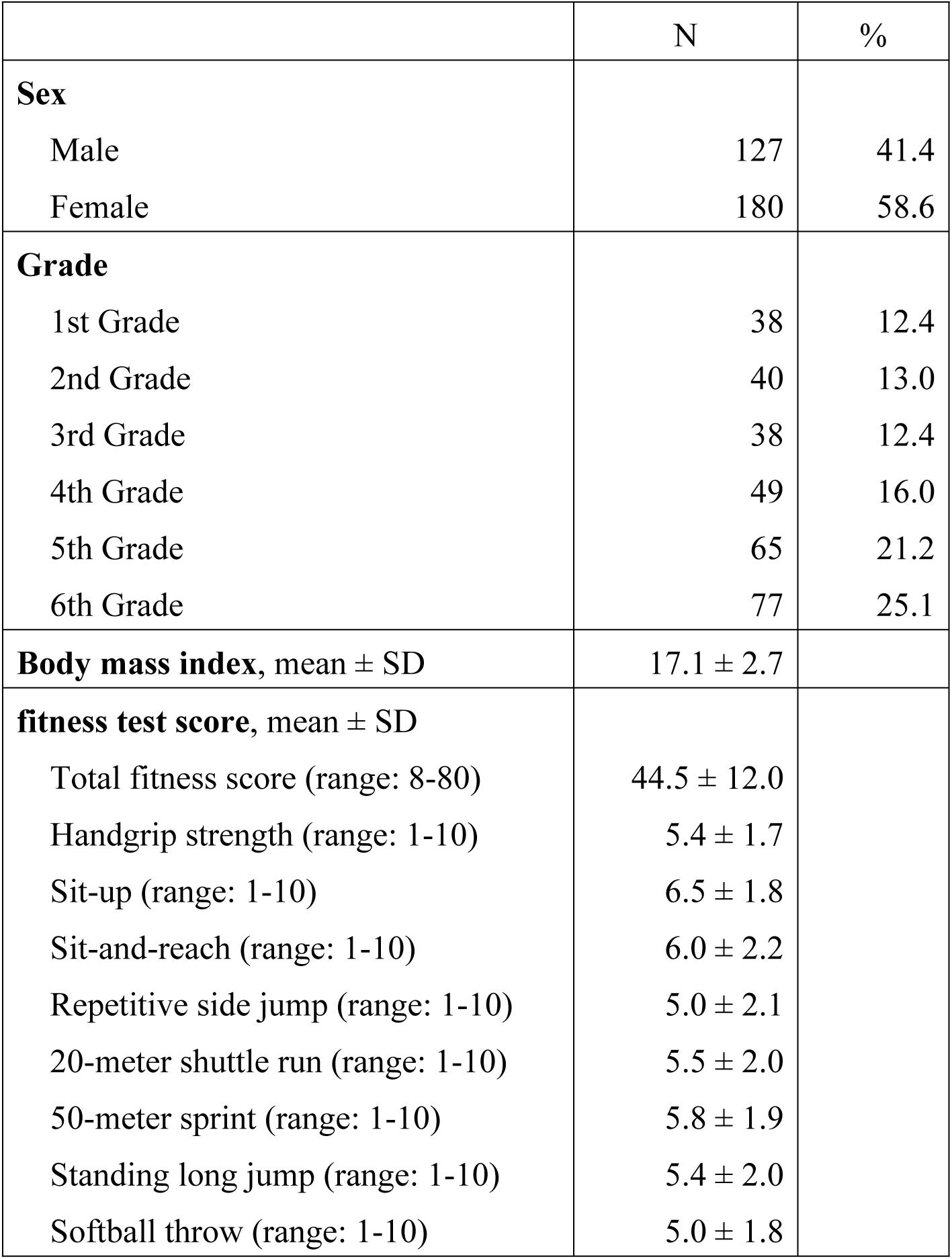
Characteristics of participants.

Table 2 presents the compliance rates for MVPA, ScT, and Sleep after 24-h MG. In this study, 4.9% (n = 15) of subjects demonstrated compliance with all three recommendations. Conversely, 25.1% (n = 77) of participants did not adhere to any of the three recommendations.

**Table 2.**
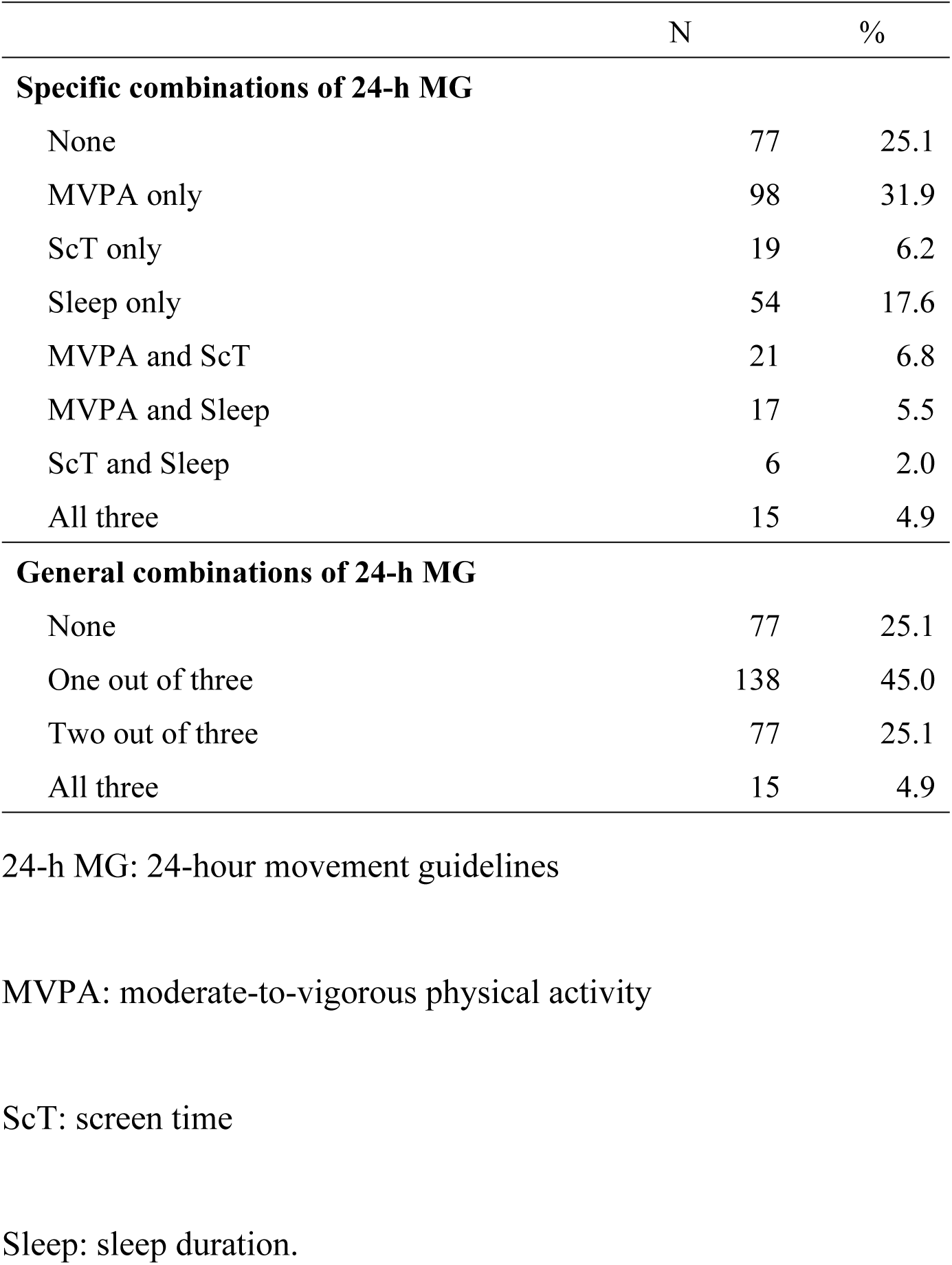
Status of Compliance with 24-h MG.

Table 3 shows the association between compliance with 24-h MG and fitness test scores. Total fitness scores were higher for only MVPA (F [1, 300] = 10.3, η2 = 0.033; P < 0.01), MVPA and ScT (F [1, 301] = 7.6, η2 = 0.025; P < 0.01), MVPA and Sleep (F [1, 301] = 11.6, η2 = 0.037; P < 0.001), and all three (F [1, 302] = 4.8, η2 = 0.016; P < 0.05) were significantly higher compared to total fitness scores when compliance for not meet.

**Table 3.**
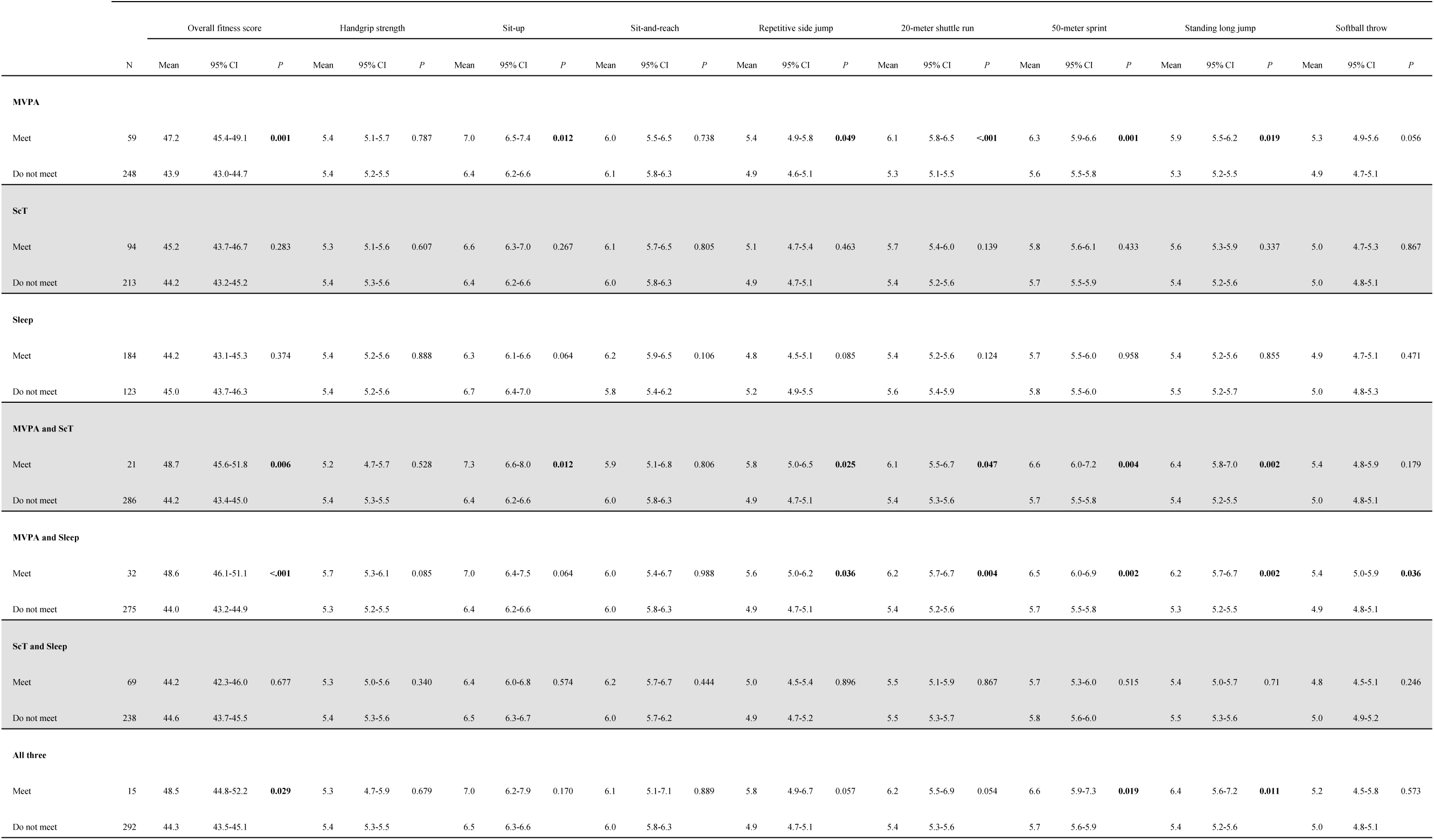
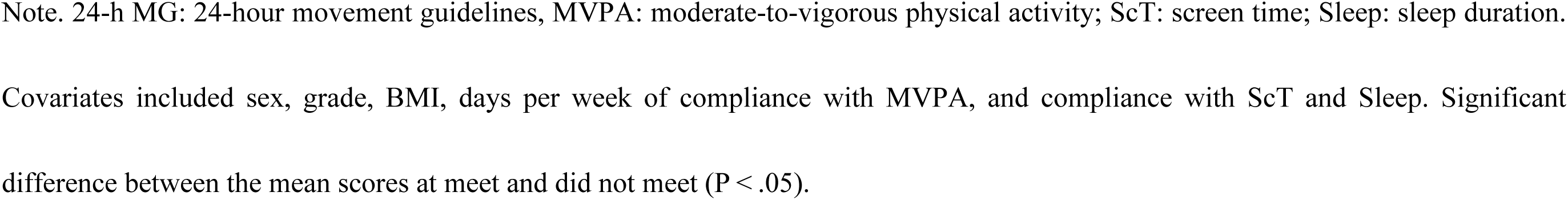
Differences in physical fitness scores on meeting patterns of 24-h MG recommendation.

For each of the physical fitness test categories, compliance with only MVPA, or MVPA and ScT, was associated with sit-up (F [1, 300] = 6.3, η2 = 0.021; P < 0.05, F [1, 301] = 6.4, η2 = 0.021; P < 0.05, respectively), repetitive side jump (F [1, 300] = 3.9, η2 = 0.013; P < 0.05, F [1, 301] = 5.1, η2 = 0.017; P < 0.05, respectively), the 20-m shuttle run (F [1, 300] = 15.9, η2 = 0.050; P < 0.001, F [1, 301] = 4.0, η2 = 0.013; P < 0.05, respectively), the 50-m sprint (F [1, 300] = 10.3, η2 = 0.033; P < 0.01, F [1, 301] = 8.4, η2 = 0.027; P < 0.01, respectively) and standing long jump (F [1, 300] = 5.6, η2 = 0.018; P < 0.05, F [1, 301] = 10.0, η2 = 0.032; P < 0.01, respectively) when compared to the students who did not comply with any component of 24-h MG. Adherence to MVPA and Sleep was associated with significantly higher scores, than non-adherence, for repetitive side jump (F [1, 301] = 4.4, η2 = 0.015; P < 0.05), 20-m shuttle run (F [1, 301] = 8.2, η2 = 0.027; P < 0.01), 50-m sprint (F [1, 301] = 9.5, η2 = 0.031; P < 0.01), standing long jump (F [1, 301] = 10.2, η2 = 0.033; P < 0.01), and softball throw (F [1, 301] = 4.5, η2 = 0.015; P < 0. 05). Meeting all three components (MVPA, ScT, and Sleep) resulted in significantly higher scores for 50-m sprint (F [1, 302] = 5.5, η2 = 0.018; P < 0.05) and standing long jump (F [1, 302] = 6.6, η2 = 0.021; P < 0.05) compared to those who did meet any of the components. Handgrip strength and sit-and-reach tests were not associated with compliance or non-compliance with any component of 24-h MG.

## Discussion

In the present study, we investigated the differences in overall physical fitness and each physical fitness component score among Japanese elementary schoolchildren in compliance with 24-h MG using standardized physical fitness test scores. Only 4.9% of children met all three criteria (MVPA, ScT, and Sleep) of 24-h MG; 25.1% of participants did not meet any of the three recommendations. The overall physical fitness score, and each physical fitness component score, were significantly higher for those who engaged in only MVPA or a combination of MVPA and other behaviors (Sleep, ScT) than for those who did not. Grip strength and flexibility were not associated with adherence to 24-h MG.

We found that the overall physical fitness score was significantly higher for those who engaged in only MVPA, MVPA and ScT, MVPA and Sleep, and all three categories. Although the number of measured events and methods were different, the results were consistent with those of a previous large-scale study[28] that investigated overall physical fitness and compliance with 24-h MG. However, in a previous study, the overall physical fitness score was significantly higher when only the ScT was performed; these findings differ from those of the present study. The factors responsible for this may include differences in the target age group, the specific items included in overall physical fitness, the proportion of subjects who met 24-h MG, and the statistical power arising from sample size. Our findings showed that overall physical fitness was significantly higher for those who met all three of 24-h MG criteria than for those who did not, thus supporting the findings of a previous study [27].

We acquired data on handgrip strength and standing long jump performance as indicators of muscle strength. As in a previous study [29], no association was observed between handgrip strength and 24-h MG. Conversely, a significant association was detected between higher scores on the standing long jump and meeting MVPA; furthermore, this association was strengthened by the combination of MVPA and ScT, and MVPA and Sleep. Despite having the same muscle strength index, the difference in our results between handgrip strength and the standing long jump may be due to a variety of reasons. First, MVPA, which includes sports activities, may have a greater influence on lower-limb muscle strength than on upper-limb muscle strength. Second, handgrip strength increases remarkably after the age of 12 years [44]. The present study was conducted in a younger age group that may have not shown an association with MVPA. A previous study in a sample of children aged 11–16 years detected a significant association between higher handgrip strength and meeting MVPA [27]. A previous study showed an association between a lower TV viewing time, which forms part of the ScT, and higher standing long jump scores; thus, this association may have been stronger in the present study with compliance to MVPA and ScT than with compliance to only MVPA.

In the present study, higher trunk flexor muscle strength and endurance (sit-up) scores were associated with meeting the only MVPA, MVPA, and ScT criteria. In a previous study [29], meeting with the only MVPA, and MVPA and Sleep, was associated with significantly higher scores for the same events; these combinations did not concur with those of the present study. While not statistically significant, the mean scores for sit-up were higher for MVPA and Sleep adherence in the present study than for those who did not; these findings were similar to those of a previous study [29]. Previous studies have demonstrated an association between a longer ScT and lower abdominal and trunk muscle strength [45]. No significant difference was demonstrated in this study between meeting with or without ScT alone; however, meeting with MVPA and ScT resulted in a significantly higher trunk flexor muscle strength and muscular endurance. Since the mean sit-up scores were higher when meeting MVPA and ScT than just for MVPA, ScT may also have some influence on the strength of the trunk flexor musculature. Previous studies have shown that prolonged exposure to ScTs, such as smartphones, games, and computers, is associated with musculoskeletal problems in the lumbar back [46,47], and that shorter ScTs may contribute to higher sit-up scores.

Next, with regards to the softball throw as a measure of instantaneous force and dexterity, significantly higher scores were observed only when meeting MVPA and Sleep requirements. Softball throwing is the most underperforming event among Japanese children when compared to scores in the 1980s (e.g., for boys in 1980: 15.4m at age 7 years, 34.0m at age 11 years; in 2021: 11.4m at age 7 years, 25.4m at age 11 years; for girls in 1980: 8.8m at age 7 years, 20.5m at age 11 years; in 2021: 7.4m at age 7 years, 15.2m at age 11 years) [44]. Although scores were significantly higher when meeting only MVPA for muscular strength indicators, such as handgrip strength, standing long jump, sit-up and softball throwing, which also require a muscular strength component, they did not show a significantly higher score when meeting only MVPA. Since softball throwing is heavily influenced by both muscle strength and dexterity, a reduction in outdoor play during childhood and early childhood may have reduced the number of opportunities to develop dexterity in coordinated body movement.

In the present study, the scores for cardiorespiratory fitness (the 20-m shuttle run) and agility (repetitive side jump) were significantly higher in those meeting only MVPA, MVPA and ScT, MVPA and Sleep, and speed (the 50-m sprint) criteria. With regards to cardiorespiratory fitness, previous studies have demonstrated an association between only MVPA[29] and MVPA and Sleep [26,27], which partially supports our present findings. Although previous studies have demonstrated an association between ScT length and lower cardiorespiratory fitness [48,49], we observed no significant differences, although cardiorespiratory fitness was higher in patients who adhered only to ScT. A previous study reported that for children and adolescents, cardiorespiratory fitness and MVPA were associated, whereas for ScT, these associations were stronger after adolescence [19]. It is possible that the association between cardiorespiratory fitness and ScT was not significant in the present study because we focused on children. A previous study[27] used a 4 × 10 m shuttle run as an indicator of agility and speed and reported significantly higher scores for those meeting only MVPA or MVPA and Sleep, thus concurring with our present results, at least in part. To the best of our knowledge, this is the only comparable study to date, and further research is now required to investigate agility and speed.

In the present study, we did not identify any significant association between flexibility (sit-and-reach) and meeting 24-h MG, thus supporting the results of a previous study[29] using a similar measurement method. However, previous studies have shown that an increased MVPA has a positive effect on flexibility [50], and more than two hours per day of ScT [51], and longer hours of computer game use [52], are negatively associated with flexibility; therefore, results are not consistent, and further investigation is required.

The present study is the first to investigate the relationship between meeting 24-h MG, overall physical fitness, and multiple components (eight events) using objective scores from standardized physical fitness tests conducted in elementary schools in Japan. Consequently, the overall physical fitness score, and many of the individual physical fitness components (core muscle strength, muscular endurance, agility, cardiorespiratory fitness, speed, and lower extremity muscle strength), were higher when meeting the MVPA alone or in combination with ScT and/or sleep. Thus, it is essential for children to meet the MVPA recommendations to enhance their physical fitness. In addition, since meeting the MVPA combined with other behaviors (ScT and Sleep) strengthened the association with overall physical fitness and some components of physical fitness (agility, lower limb muscle strength, and skillfulness) when compared with the MVPA alone, compliance with ScT and Sleep criteria may contribute to enhancing physical fitness in students attending elementary schools.

The present study has several limitations that need to be considered. First, the MVPA, ScT, and Sleep data were collected using questionnaires, which may be subject to recall bias. Second, owing to missing response data, only 65% of the total dataset were included in our final analysis. Third, the survey was conducted at a single school. Finally, since this was a cross-sectional study, the causal relationships between compliance with 24-hour MG and physical fitness remain unclear.

## Conclusion

In conclusion, meeting MVPA criteria was strongly associated with higher overall fitness and physical fitness components in children. Furthermore, compliance with MVPA combined with ScT and/or sleep criteria strengthened these relationships.

## Data Availability

The datasets generated and/or analyzed in the current study are not publicly available due to ethical considerations however, they are available from the corresponding author upon reasonable request.

## Acknowledgements

We would like to express our appreciation to all the children, their parents, and teachers who participated in the research. This study was supported by JSPS KAKENHI (Grant number: JP 21K11507, JP23K10770).

## Notes

### Competing Interest Statement

The authors have declared no competing interest.

### Funding Statement

Yes

### Author Declarations

Waseda University Ethics Review Procedures Concerning Research with Human Subjects

## References

1. Guthold R, Stevens GA, Riley LM, Bull FC. Global trends in insufficient physical activity among adolescents: a pooled analysis of 298 population-based surveys with 1·6 million participants. Lancet Child Adolesc Health. 2020;4: 23–35. doi:10.1016/s2352-4642(19)30323-2

2. Yang L, Cao C, Kantor ED, Nguyen LH, Zheng X, Park Y, et al. Trends in Sedentary Behavior Among the US Population, 2001-2016. JAMA. 2019;321: 1587–1597. doi:10.1001/jama.2019.3636

3. Aubert S, Barnes JD, Demchenko I, Hawthorne M, Abdeta C, Abi Nader P, et al. Global Matrix 4.0 Physical Activity Report Card Grades for Children and Adolescents: Results and Analyses From 57 Countries. J Phys Act Health. 2022;19: 700–728. doi:10.1123/jpah.2022-0456

4. Lamoureux NR, Fitzgerald JS, Norton KI, Sabato T, Tremblay MS, Tomkinson GR. Temporal Trends in the Cardiorespiratory Fitness of 2,525,827 Adults Between 1967 and 2016: A Systematic Review. Sports Med. 2019;49: 41–55. doi:10.1007/s40279-018-1017-y

5. Tomkinson GR, Lang JJ, Tremblay MS. Temporal trends in the cardiorespiratory fitness of children and adolescents representing 19 high-income and upper middle-income countries between 1981 and 2014. Br J Sports Med. 2019;53: 478–486. doi:10.1136/bjsports-2017-097982

6. Sandercock GRH, Cohen DD. Temporal trends in muscular fitness of English 10-year-olds 1998-2014: An allometric approach. J Sci Med Sport. 2019;22: 201–205. doi:10.1016/j.jsams.2018.07.020

7. Masanovic B, Gardasevic J, Marques A, Peralta M, Demetriou Y, Sturm DJ, et al. Trends in Physical Fitness Among School-Aged Children and Adolescents: A Systematic Review. Front Pediatr. 2020;8: 627529. doi:10.3389/fped.2020.627529

8. Tomkinson GR, Kaster T, Dooley FL, Fitzgerald JS, Annandale M, Ferrar K, et al. Temporal Trends in the Standing Broad Jump Performance of 10,940,801 Children and Adolescents Between 1960 and 2017. Sports Med. 2021;51: 531–548. doi:10.1007/s40279-020-01394-6

9. Nishijima T, Kokudo S, Ohsawa S. Changes over the years in physical and motor ability in Japanese youth in 1964-97. Int J Sport Health Sci. 2003;1: 164–170. doi:10.5432/ijshs.1.164

10. Japan Sports Agency. Report on the Physical Fitness and Exercise Capacity Survey in 2018. 2019 [cited 1 Nov 2024]. Available: https://www.mext.go.jp/sports/b_menu/toukei/chousa04/tairyoku/kekka/k_detail/1421920.htm (in Japanese)

11. Central Council for Education. Comprehensive strategies to improve children’s physical fitness. In: Ministry of Education, Culture, Sports, Science and Technology [Internet]. 2002 [cited 1 Nov 2024]. Available: https://www.mext.go.jp/b_menu/shingi/chukyo/chukyo0/toushin/021001.htm (in Japanese)

12. Yogi Y, Ishikawa Y, Takahashi S. Secular Contrasts in Physical Fitness and Athletic Skills in Japanese Elementary School Students (11-Year-Olds). Int J Environ Res Public Health. 2024;21. doi:10.3390/ijerph21070951

13. Janssen I, Leblanc AG. Systematic review of the health benefits of physical activity and fitness in school-aged children and youth. Int J Behav Nutr Phys Act. 2010;7: 40. doi:10.1186/1479-5868-7-40

14. Smith JJ, Eather N, Morgan PJ, Plotnikoff RC, Faigenbaum AD, Lubans DR. The health benefits of muscular fitness for children and adolescents: a systematic review and meta-analysis. Sports Med. 2014;44: 1209–1223. doi:10.1007/s40279-014-0196-4

15. Haapala EA, Leppänen MH, Skog H, Lubans DR, Viitasalo A, Lintu N, et al. Childhood Physical Fitness as a Predictor of Cognition and Mental Health in Adolescence: The PANIC Study. Sports Med. 2024. doi:10.1007/s40279-024-02107-z

16. Tremblay MS, LeBlanc AG, Kho ME, Saunders TJ, Larouche R, Colley RC, et al. Systematic review of sedentary behaviour and health indicators in school-aged children and youth. Int J Behav Nutr Phys Act. 2011;8: 98. doi:10.1186/1479-5868-8-98

17. Carson V, Hunter S, Kuzik N, Gray CE, Poitras VJ, Chaput JP, et al. Systematic review of sedentary behaviour and health indicators in school-aged children and youth: an update. Appl Physiol Nutr Metab. 2016;41: S240–65. doi:10.1139/apnm-2015-0630

18. Fonseca A, de Azevedo CVM, Santos RMR. Sleep and health-related physical fitness in children and adolescents: a systematic review. Sleep Sci. 2021;14: 357–365. doi:10.5935/1984-0063.20200125

19. Wilhite K, Booker B, Huang BH, Antczak D, Corbett L, Parker P, et al. Combinations of Physical Activity, Sedentary Behavior, and Sleep Duration and Their Associations With Physical, Psychological, and Educational Outcomes in Children and Adolescents: A Systematic Review. Am J Epidemiol. 2023;192: 665–679. doi:10.1093/aje/kwac212

20. Tremblay MS, Carson V, Chaput JP, Connor Gorber S, Dinh T, Duggan M, et al. Canadian 24-Hour Movement Guidelines for Children and Youth: An Integration of Physical Activity, Sedentary Behaviour, and Sleep. Appl Physiol Nutr Metab. 2016;41: S311–27. doi:10.1139/apnm-2016-0151

21. Tremblay MS, Chaput JP, Adamo KB, Aubert S, Barnes JD, Choquette L, et al. Canadian 24-Hour Movement Guidelines for the Early Years (0-4 years): An Integration of Physical Activity, Sedentary Behaviour, and Sleep. BMC Public Health. 2017;17: 874. doi:10.1186/s12889-017-4859-6

22. Australian Government Department of Health. Physical activity and exercise guidelines for all Australians. In: Australian Government Department of Health and Aged Care [Internet]. 2019 [cited 1 Nov 2024]. Available: https://www.health.gov.au/topics/physical-activity-and-exercise/physical-activity-and-exercise-guidelines-for-all-australians

23. World Health Organization. Guidelines on physical activity, sedentary behaviour and sleep for children under 5 years of age. In: World Health Organization [Internet]. World Health Organization; 2 Apr 2019 [cited 1 Nov 2024]. Available: https://www.who.int/publications/i/item/9789241550536

24. Tomaz SA, Okely AD, van Heerden A, Vilakazi K, Samuels ML, Draper CE. The South African 24-Hour Movement Guidelines for Birth to 5 Years: Results From the Stakeholder Consultation. J Phys Act Health. 2020;17: 126–137. doi:10.1123/jpah.2019-0188

25. Australian Government Department of Health, Care A. Physical activity and exercise guidelines For children and young people (5 to 17 years). In: Australian Government Department of Health and Aged Care [Internet]. 2019 [cited 1 Nov 2024]. Available: https://www.health.gov.au/topics/physical-activity-and-exercise/physical-activity-and-exercise-guidelines-for-all-australians/for-children-and-young-people-5-to-17-years

26. Carson V, Chaput JP, Janssen I, Tremblay MS. Health associations with meeting new 24-hour movement guidelines for Canadian children and youth. Prev Med. 2017;95: 7–13. doi:10.1016/j.ypmed.2016.12.005

27. Tapia-Serrano MÁ, López-Gil JF, Sevil-Serrano J, García-Hermoso A, Sánchez-Miguel PA. What is the role of adherence to 24-hour movement guidelines in relation to physical fitness components among adolescents? Scand J Med Sci Sports. 2023;33: 1373–1383. doi:10.1111/sms.14357

28. Cai S, Zhong P, Dang J, Liu Y, Shi D, Chen Z, et al. Associations between combinations of 24-h movement behaviors and physical fitness among Chinese adolescents: Sex and age disparities. Scand J Med Sci Sports. 2023;33: 1779–1791. doi:10.1111/sms.14427

29. Tanaka C, Tremblay MS, Okuda M, Tanaka S. Association between 24-hour movement guidelines and physical fitness in children. Pediatr Int. 2020;62: 1381–1387. doi:10.1111/ped.14322

30. Zhao H, Wu N, Haapala EA, Gao Y. Association between meeting 24-h movement guidelines and health in children and adolescents aged 5-17 years: a systematic review and meta-analysis. Front Public Health. 2024;12: 1351972. doi:10.3389/fpubh.2024.1351972

31. Roman-Viñas B, Chaput J-P, Katzmarzyk PT, Fogelholm M, Lambert EV, Maher C, et al. Proportion of children meeting recommendations for 24-hour movement guidelines and associations with adiposity in a 12-country study. Int J Behav Nutr Phys Act. 2016;13: 123. doi:10.1186/s12966-016-0449-8

32. Jakubec L, Gába A, Dygrýn J, Rubín L, Šimůnek A, Sigmund E. Is adherence to the 24-hour movement guidelines associated with a reduced risk of adiposity among children and adolescents? BMC Public Health. 2020;20: 1119. doi:10.1186/s12889-020-09213-3

33. Fairclough SJ, Clifford L, Brown D, Tyler R. Characteristics of 24-hour movement behaviours and their associations with mental health in children and adolescents. J Act Sedentary Sleep Behav. 2023;2: 11. doi:10.1186/s44167-023-00021-9

34. Sampasa-Kanyinga H, Colman I, Goldfield GS, Janssen I, Wang J, Podinic I, et al. Combinations of physical activity, sedentary time, and sleep duration and their associations with depressive symptoms and other mental health problems in children and adolescents: a systematic review. Int J Behav Nutr Phys Act. 2020;17: 72. doi:10.1186/s12966-020-00976-x

35. Kyan A, Takakura M, Miyagi M. Associations between 24-h movement behaviors and self-rated health: a representative sample of school-aged children and adolescents in Okinawa, Japan. Public Health. 2022;213: 117–123. doi:10.1016/j.puhe.2022.10.012

36. Fung H, Yeo BTT, Chen C, Lo JC, Chee MWL, Ong JL. Adherence to 24-Hour Movement Recommendations and Health Indicators in Early Adolescence: Cross-Sectional and Longitudinal Associations in the Adolescent Brain Cognitive Development Study. J Adolesc Health. 2023;72: 460–470. doi:10.1016/j.jadohealth.2022.10.019

37. Watson A, Dumuid D, Maher C, Olds T. Associations between meeting 24-hour movement guidelines and academic achievement in Australian primary school-aged children. J Sport Health Sci. 2022;11: 521–529. doi:10.1016/j.jshs.2020.12.004

38. Lien A, Sampasa-Kanyinga H, Colman I, Hamilton HA, Chaput J-P. Adherence to 24-hour movement guidelines and academic performance in adolescents. Public Health. 2020;183: 8–14. doi:10.1016/j.puhe.2020.03.011

39. Tapia-Serrano MA, García-Hermoso A, Sevil-Serrano J, Sánchez-Oliva D, Sánchez-Miguel PA. Is adherence to 24-Hour Movement Guidelines associated with a higher academic achievement among adolescent males and females? J Sci Med Sport. 2022;25: 155–161. doi:10.1016/j.jsams.2021.09.005

40. Ministry of Education, Culture, Sports, Science and Technology. New Physical Fitness Test Implementation Guidelines. In: Ministry of Education, Culture, Sports, Science and Technology [Internet]. 1999 [cited 1 Nov 2024]. Available: https://www.mext.go.jp/sports/b_menu/sports/mcatetop03/list/detail/1408001.htm (in Japanese)

41. Janssen I, Katzmarzyk PT, Boyce WF, King MA, Pickett W. Overweight and obesity in Canadian adolescents and their associations with dietary habits and physical activity patterns. J Adolesc Health. 2004;35: 360–367. doi:10.1016/j.jadohealth.2003.11.095

42. Ishii K, Shibata A, Adachi M, Mano Y, Oka K. School grade and sex differences in domain-specific sedentary behaviors among Japanese elementary school children: a cross-sectional study. BMC Public Health. 2017;17: 318. doi:10.1186/s12889-017-4221-z

43. Japanese Society of School Health. Surveillance Report on Children’s Health. 2010. In: Japanese Society of School Health [Internet]. 2010 [cited 1 Nov 2024]. Available: http://www.gakkohoken.jp/book/ebook/ebook_H250060/index_h5.html (in Japanese)

44. Japan Sports Agency. Results of the Physical Fitness and Motor Skills Survey in 2022. In: Japan Sports Agency [Internet]. 2023 [cited 1 Nov 2024]. Available: https://www.mext.go.jp/sports/b_menu/toukei/chousa04/tairyoku/kekka/k_detail/1421920_00010.htm (in Japanese)

45. Grøntved A, Ried-Larsen M, Froberg K, Wedderkopp N, Brage S, Kristensen PL, et al. Screen time viewing behaviors and isometric trunk muscle strength in youth. Med Sci Sports Exerc. 2013;45: 1975–1980. doi:10.1249/MSS.0b013e318295af56

46. Cankurtaran F, Menevşe O, Namlı A, Kızıltoprak HŞ, Altay S, Duran M, et al. The impact of digital game addiction on musculoskeletal system of secondary school children. Niger J Clin Pract. 2022;25: 153–159. doi:10.4103/njcp.njcp_177_20

47. Azevedo N, Ribeiro JC, Machado L. Back pain in children and adolescents: a cross-sectional study. Eur Spine J. 2023;32: 3280–3289. doi:10.1007/s00586-023-07751-z

48. Mitchell JA, Pate RR, Blair SN. Screen-based sedentary behavior and cardiorespiratory fitness from age 11 to 13. Med Sci Sports Exerc. 2012;44: 1302–1309. doi:10.1249/MSS.0b013e318247cd73

49. Sandercock GRH, Ogunleye AA. Independence of physical activity and screen time as predictors of cardiorespiratory fitness in youth. Pediatr Res. 2013;73: 692–697. doi:10.1038/pr.2013.37

50. Aggio D, Smith L, Hamer M. Effects of reallocating time in different activity intensities on health and fitness: a cross sectional study. Int J Behav Nutr Phys Act. 2015;12: 83. doi:10.1186/s12966-015-0249-6

51. Tucker JS, Martin S, Jackson AW, Morrow JR Jr, Greenleaf CA, Petrie TA. Relations between sedentary behavior and FITNESSGRAM healthy fitness zone achievement and physical activity. J Phys Act Health. 2014;11: 1006–1011. doi:10.1123/jpah.2011-0431

52. Cieśla E, Mleczko E, Bergier J, Markowska M, Nowak-Starz G. Health-Related Physical Fitness, BMI, physical activity and time spent at a computer screen in 6 and 7-year-old children from rural areas in Poland. Ann Agric Environ Med. 2014;21: 617–621. doi:10.5604/12321966.1120613

